# Public awareness of the hearing loss-dementia link and self-reported behavioral changes in middle-to older-aged Canadians

**DOI:** 10.64898/2026.01.12.26343953

**Authors:** Sarah Bobbitt, Signe Lund Mathiesen, Malcolm Binns, Björn Herrmann

**Author notes:** Correspondence concerning this article should be addressed to Björn Herrmann, Rotman Research Institute, Baycrest, 3560 Bathurst St, North York, ON, M6A 2E1, Canada. **Conflict of interest statement:** The authors declare no competing interests.

## Abstract

Hearing loss is a potential risk factor for dementia, but public awareness of this link and its behavioral implications remain poorly understood. In a cross-sectional survey, 1,404 Canadians aged 44-97 years completed measures of awareness, subjective hearing and cognition, dementia-related concerns, perceptions of the link, and demographics. 28% of people were aware of the hearing loss–dementia link. Of these, 17% were more inclined to seek/continue hearing-loss treatment. Awareness and self-reported behavioral responses were over twice as prevalent in a health-engaged subsample. Education, dementia concern, older age, and female gender were associated with awareness. Among aware participants, dementia concern, poorer subjective cognition, and confidence in understanding the link, but not education, were associated with reported treatment inclination. Awareness thus follows socioeconomic and demographic gradients, but these alone do not appear to ensure behavior change. Communication may need to reach beyond health-engaged groups and prioritize accurate understanding over awareness.

## Introduction

Dementia research has increasingly emphasized prevention through modifiable risk factors. The Lancet Commission on dementia prevention, intervention, and care estimated that about 45% of dementia cases worldwide are attributable to 14 potentially modifiable risk factors (Livingston et al., 2024), and informing the public about these factors has become a component of dementia-prevention efforts (Van Asbroeck et al., 2021; Paauw et al., 2024; Eddy-Lacey et al., 2025). Hearing loss has been among these factors since the Commission’s first reports (Livingston et al., 2017; Livingston et al., 2020). Age-related hearing loss affects about 40% of adults aged 65 years and older (Cruickshanks et al., 1998; Feder et al., 2015; Goman and Lin, 2016), and longitudinal studies have associated it with accelerated cognitive decline and increased dementia incidence (Lin et al., 2011; Chern and Golub, 2019; Panza et al., 2019; Park et al., 2022; Son et al., 2024), even after adjustment for demographic and health factors (Lin et al., 2013; Chern and Golub, 2019; Croll et al., 2021; Kolo et al., 2025).

The link between hearing loss and dementia has since been communicated widely beyond the scientific literature. The Lancet Commission reports received extensive media coverage (Timson, 2017; Jiang, 2024b; Pope, 2024), news outlets have described hearing loss as a leading modifiable risk factor for dementia (Taylor, 2018; McKnight, 2019; La Grassa, 2023; Jiang, 2024a), and hearing-care providers refer to dementia prevention in patient-facing materials (Blustein et al., 2020; Wix, 2024; Herrmann et al., 2026). By framing hearing care as a way to protect cognition, such messages may encourage people to seek hearing assessment or to adopt hearing aids. This potential is considerable because hearing loss in older adults often goes untreated, and only a minority of adults who could benefit from hearing aids use them, typically after delays of many years (Chien and Lin, 2013; Bisgaard and Ruf, 2017; Simpson et al., 2019).

Whether treating hearing loss slows cognitive decline, however, remains unresolved. Observational and intervention studies have yielded mixed results (Maharani et al., 2018; Sarant et al., 2020; Sanders et al., 2021), and a large, randomized trial found slower cognitive decline only in a subgroup at elevated risk due to age or comorbidities (Lin et al., 2023), with modest effect sizes at the individual level (Dawes and Munro, 2024; Sarant et al., 2024). Messages that link hearing care to dementia prevention may thus create expectations of cognitive benefits that might exceed the current evidence (Blustein et al., 2020; Pichora-Fuller, 2023), and they may add to worries about dementia, which are common among older adults (Molden and Maxfield, 2017; Werner et al., 2021). Whether such communication does more good than harm thus depends on who encounters it and on how they interpret and act on it.

Little is known about who has encountered the message linking hearing loss to dementia or whether they change behavior as a result of know about the link. Knowledge of modifiable dementia risk factors in the general population is limited, with about 40 to 50% of adults able to identify at least one risk factor (Cations et al., 2018; Heger et al., 2019; Zheng et al., 2020). In surveys that included hearing loss among candidate risk factors, 4 to 35% of respondents in Australia, Iceland, Ireland, and Norway identified it (Nagel et al., 2021; Jónsdóttir et al., 2022; Kjelvik et al., 2022; Dukelow et al., 2023), compared with 59% in Bulgaria (Lazarova and Petrova-Antonova, 2025). However, these surveys assessed dementia knowledge in general. They did not examine who has encountered the hearing loss–dementia link, whether people are aware of claims that hearing aids may benefit cognition, or whether this information has influenced hearing-related behavior. The last question is important because awareness of a health risk is often insufficient to change behavior (Marteau et al., 2012; Kelly and Barker, 2016).

The COM-B model of behavior (Michie et al., 2011), previously applied to hearing-aid use (Barker et al., 2016), can structure these questions. It proposes that behavior arises from capability (e.g., knowledge and understanding), opportunity (e.g., access to information and services), and motivation (e.g., perceived relevance, worry, and trust). We thus distinguish between the awareness of the hearing loss–dementia link, which should depend mainly on opportunities to encounter health information (e.g., education, income, age, gender, and place of residence), and the response to it, which should depend more on capability and motivation to act (e.g., subjective hearing and cognitive difficulties, dementia concern, confidence in understanding the link, and trust in healthcare professionals).

Canada provides a relevant context for examining these questions. Its population is aging rapidly (Statistics-Canada, 2022), and Canadian news media have repeatedly covered the link between hearing loss and dementia over the past decade (Taylor, 2018; McKnight, 2019; La Grassa, 2023; Jiang, 2024a). Hearing care is funded through a mix of public and private systems, and coverage of hearing assessments and hearing aids differs across provinces (Fournier, 2017), which may shape both the opportunity to encounter hearing-related health information and the capacity to act on it. Moreover, none of the awareness surveys described above was conducted in Canada. The present study examined the awareness of, and self-reported behavioral responses to, the hearing loss–dementia message among middle-aged and older Canadians. Aim 1 (awareness) was to estimate how many people have heard about (i) a possible link between hearing loss and cognitive decline/dementia and (ii) the claim that hearing aids might help to prevent or reduce cognitive decline/dementia, and to identify characteristics associated with having heard these messages. Aim 2 (response) was to estimate, among those who had heard about the link, how many report that it made them (i) more inclined to seek or continue hearing-loss treatment or (ii) more open in talking about their hearing loss, and to identify characteristics associated with these self-reported responses.

## Methods and Materials

### Study Design, Recruitment, and Participants

This cross-sectional online survey was administered through Qualtrics from May to August 2025. Participants had to reside in Canada and be native or proficient English speakers. Recruitment used two commercial panels, Canadian Viewpoint (https://canview.com/) and Decision Point Research (https://decisionpointresearch.ca/), and newsletters or social media associated with Cogniciti, the Canadian Hard of Hearing Association, AGE-WELL Network, and the Canadian Association of Retired Persons. Recruitment through Canadian Viewpoint was larger because it was more cost-effective. Participants received direct compensation or entry into a raffle. Of 1884 recorded responses, 480 were excluded due to eligibility, consent, English proficiency, incomplete responses, completion time, and attention checks (Supplementary Methods). The final sample comprised 1404 participants (Canadian Viewpoint: 948; Decision Point: 170; convenience sample: 286; age range: 44–97 years, median age: 68 years, 784 females, 612 males; no participant identified with any other gender; 8 did not report their gender).

### Ethics declaration

Participants gave informed consent prior to the study by selecting “Yes, I agree to take part in this research” in the online form. The study was conducted in accordance with the Declaration of Helsinki and the Canadian Tri-Council Policy Statement on Ethical Conduct for Research Involving Humans (TCPS2-2014), and was approved by the Research Ethics Board of the Rotman Research Institute at Baycrest Academy for Research and Education.

### Survey Measures

The survey included items assessing knowledge and perceptions of the link between hearing loss and cognitive decline/dementia, subjective hearing abilities, subjective cognitive abilities and concerns, and demographic characteristics (e.g., age, gender, education, household income, postal code), as well as attention checks (Supplementary Materials). The variables analyzed are listed in Table 1. The survey is provided on https://osf.io/f4d9t/ (publicly available upon publication).

**Table 1:**
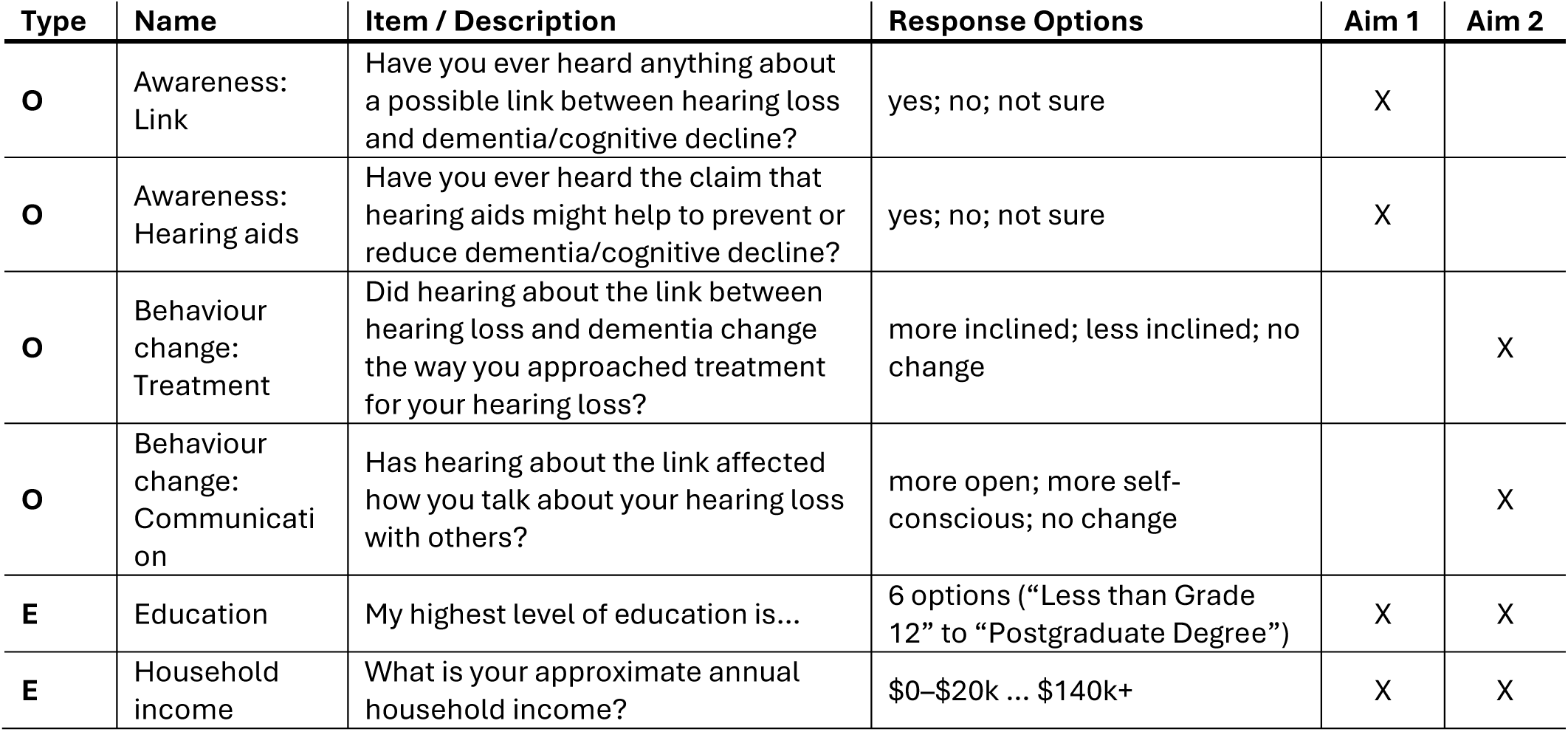
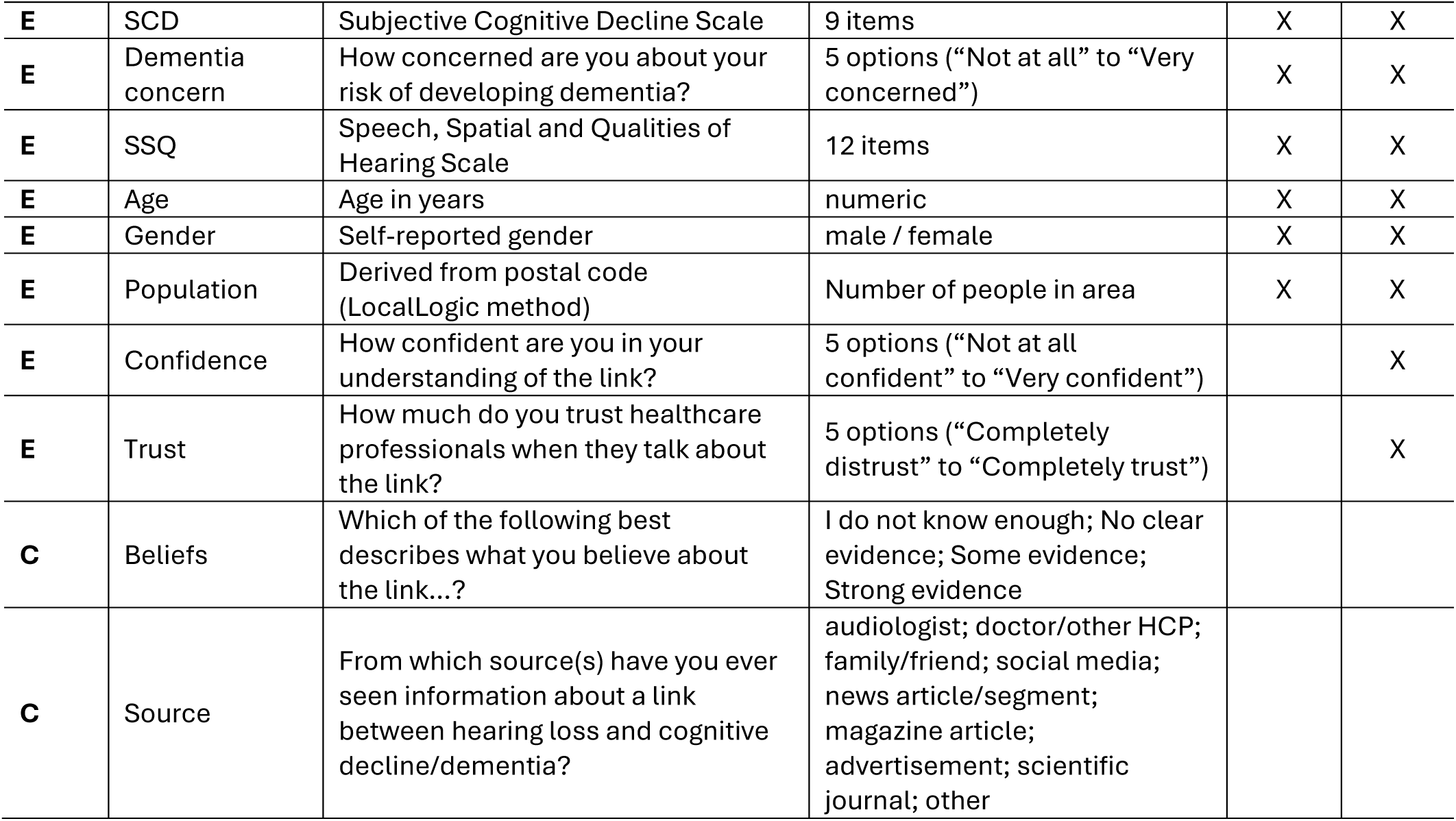
Variables used in the current study. Outcome variable, P – Explanatory variable, C – Context variable

| Type | Name | Item / Description | Response Options | Aim 1 | Aim 2 |
| --- | --- | --- | --- | --- | --- |
| O | Awareness: Link | Have you ever heard anything about a possible link between hearing loss and dementia/cognitive decline? | yes; no; not sure | X |  |
| O | Awareness: Hearing aids | Have you ever heard the claim that hearing aids might help to prevent or reduce dementia/cognitive decline? | yes; no; not sure | X |  |
| O | Behaviour change: Treatment | Did hearing about the link between hearing loss and dementia change the way you approached treatment for your hearing loss? | more inclined; less inclined; no change |  | X |
| O | Behaviour change: Communication | Has hearing about the link affected how you talk about your hearing loss with others? | more open; more self-conscious; no change |  | X |
| E | Education | My highest level of education is... | 6 options ("Less than Grade 12" to "Postgraduate Degree") | X | X |
| E | Household income | What is your approximate annual household income? | \$0–\$20k ... \$140k+ | X | X |
| **E** | SCD | Subjective Cognitive Decline Scale | 9 items | X | X |
| **E** | Dementia concern | How concerned are you about your risk of developing dementia? | 5 options (“Not at all” to “Very concerned”) | X | X |
| **E** | SSQ | Speech, Spatial and Qualities of Hearing Scale | 12 items | X | X |
| **E** | Age | Age in years | numeric | X | X |
| **E** | Gender | Self-reported gender | male / female | X | X |
| **E** | Population | Derived from postal code (LocalLogic method) | Number of people in area | X | X |
| **E** | Confidence | How confident are you in your understanding of the link? | 5 options (“Not at all confident” to “Very confident”) |  | X |
| **E** | Trust | How much do you trust healthcare professionals when they talk about the link? | 5 options (“Completely distrust” to “Completely trust”) |  | X |
| **C** | Beliefs | Which of the following best describes what you believe about the link...? | I do not know enough; No clear evidence; Some evidence; Strong evidence |  |  |
| **C** | Source | From which source(s) have you ever seen information about a link between hearing loss and cognitive decline/dementia? | audiologist; doctor/other HCP; family/friend; social media; news article/segment; magazine article; advertisement; scientific journal; other |  |  |

#### Outcome Variables for Aim 1: Awareness of the Link

Two outcome items assessed awareness of the hearing loss–dementia link:

1. *“Have you ever heard anything about a possible link between hearing loss and dementia/cognitive decline (e.g., a decline in memory, attention, problem solving, etc.)?”* (response options: “yes”, “no”, “not sure”).
2. *“Have you ever heard the claim that hearing aids might help to prevent or reduce dementia/cognitive decline?”* (response options: “yes”, “no”, “not sure”).

#### Outcome Variables for Aim 2: Self-Reported Behavioral Response

Participants who had heard about the link were asked two further questions about their perceived response to it:

1. *“Did hearing about the link between hearing loss and dementia change the way you approached treatment for your hearing loss?”* (response options: “Yes, it made me more inclined to seek/continue treatment”, “Yes, it made me less inclined to seek/continue treatment,” “No, it did not change anything”).
2. *“Has hearing about the link between hearing loss and cognitive decline affected how you talk about your hearing loss with others (e.g., family, friends, healthcare providers)?”*

(response options: “Yes, I talk about it more openly”, “Yes, but I now feel more self-conscious or worried”, “No, it has not changed how I talk about it”).

#### Explanatory Variables

The survey was not designed around a formal behavioral model, but explanatory variables were grouped according to the COM-B framework outlined in the Introduction (Michie et al., 2011). Variables expected to shape opportunities to encounter, and capability to process, health information were education (*My highest level of education is…*; six levels from “Less than Grade 12” to “Postgraduate Degree”), household income (*What is your approximate annual household income (before taxes)?*; eight levels from “$0–$20,000” to “$140,000+”; pre-retirement income for retirees), age, gender, and population size. Population size, a proxy for urbanicity, was derived for each participant’s postal code using Local Logic data (https://locallogic.co/), which estimates the number of people living in an area. Variables expected to shape personal relevance and motivation were subjective hearing abilities (12-item Speech, Spatial and Qualities of Hearing scale, SSQ; Gatehouse and Noble, 2004; Noble et al., 2013), subjective cognitive decline (9-item SCD; Gifford et al., 2015), and dementia concern (*How concerned are you about your risk of developing dementia?*; five levels from “Not at all concerned” to “Very concerned”).

Two additional variables, asked only of participants who had heard about the link and therefore included only in Aim 2 analyses, captured perceptions of the message itself: confidence in understanding the link (*How confident are you in your understanding of the link between hearing loss and cognitive decline/dementia?*; five levels from “Not at all confident” to “Very confident”) and trust in healthcare professionals (*How much do you trust healthcare professionals when they talk about the link between hearing loss and cognitive decline?*; five levels from “Completely distrust” to “Completely trust”).

#### Context Variables

Two additional survey items were summarized descriptively rather than modeled. The first assessed participants’ *beliefs about the link* (*“Which of the following best describes what you believe about the link between hearing loss and dementia?”*; Response options: “I do not know enough to say”, “There is no clear scientific evidence of a link between hearing loss and dementia”, “There is some scientific evidence that hearing loss increases the risk of dementia”, “There is strong scientific evidence that hearing loss causes dementia”). Because the response categories are qualitatively distinct and the item overlapped with confidence in understanding the link, confidence was used in the regression models instead. The second item asked from which source(s) participants had seen information about the link (audiologist; doctor or other healthcare professional; family member/friend; social media; news article/segment; magazine article; advertisement; scientific journal; other). This variable has nominal, non-ordinal categories, and because participants could identify multiple sources, its model inclusion is not straightforward. Both context variables were thus summarized for descriptive purposes.

### Analytic Sample

Primary regression analyses used the Canadian Viewpoint sample, the largest sample and the one with approximately equal proportions of female and male respondents. Restricting the primary models to a single recruitment source also avoids the added complexity of modeling source effects. The convenience sample was recruited through health and aging networks and is termed “health-engaged” as shorthand for its recruitment source, rather than a measured characteristic. It differed from the panels on several demographic and subjective health measures and had higher awareness (Table 2). These differences motivated separate descriptive reporting by source. Although an online panel is not a probability sample of the Canadian population, it is less selected for health engagement than the “health-engaged” convenience sample. Supplementary models that include all three samples, with recruitment source as a covariate, mirror and in places extend the results reported here (Tables S1–S4). Any divergence is noted in the Results.

**Table 2:** Descriptive data and comparisons between different recruitment sources. The three columns on the left reflect data for relevant survey variables for the three recruitment sources. Age, education, income, SCD, SSQ, and dementia concern reflect the mean across participants. Gender reflects the proportion of female. For the two awareness and two self-reported behavior responses, the proportion of ‘yes’ responses is provided. The three columns on the right side reflect significance levels for contrasts between the different recruitment sources using a t-test (continuous) or chi-square test (categorical). Corresponding effect sizes Cohen’s d and Cramer’s V are provided in parentheses for continuous and categorical variables, respectively; CV – Canadian Viewpoint, DP – Decision Point, HE – Health-Engaged.

| Variable | Canadian Viewpoint | Decision Point | Health-Engaged | CV vs DP | CV vs HE | DP vs HE |
| --- | --- | --- | --- | --- | --- | --- |
| N | 948 | 170 | 286 | - | - | - |
| Population | 1246 | 1584 | 1643 | **0.026 (0.191)** | **0.002 (0.213)** | 0.788 (0.027) |
| Age | 66.85 | 68.22 | 72.35 | 0.054 (0.162) | **<0.001 (0.620)** | **<0.001 (0.520)** |
| Gender | 0.503 | 0.621 | 0.722 | **0.005 (0.085)** | **<0.001 (0.185)** | **0.025 (0.105)** |
| Education | 3.842 | 4.170 | 4.753 | **0.007 (0.229)** | **<0.001 (0.654)** | **<0.001 (0.436)** |
| Income | 4.259 | 4.635 | 5.430 | **0.030 (0.182)** | **<0.001 (0.581)** | **<0.001 (0.390)** |
| SCD | 2.670 | 2.262 | 3.056 | **0.029 (0.182)** | **0.030 (0.147)** | **<0.001 (0.336)** |
| SSQ | 6.930 | 7.011 | 6.512 | 0.577 (0.046) | **0.001 (0.220)** | **0.014 (0.240)** |
| Dementia concern | 0.126 | 0.112 | 0.497 | 0.887 (0.012) | **<0.001 (0.327)** | **<0.001 (0.344)** |
| Awareness: Link | 0.275 | 0.359 | 0.804 | **0.027 (0.066)** | **<0.001 (0.456)** | **<0.001 (0.448)** |
| Awareness: Hearing aids | 0.134 | 0.253 | 0.615 | **<0.001 (0.119)** | **<0.001 (0.472)** | **<0.001 (0.351)** |
| Reported response: Treatment | 0.169 | 0.279 | 0.417 | **0.048 (0.110)** | **<0.001 (0.275)** | **0.048 (0.116)** |
| Reported response: Communication | 0.123 | 0.262 | 0.300 | **0.006 (0.154)** | **<0.001 (0.219)** | **0.565 (0.034)** |

## Data Analysis

For Aim 1, we fitted two logistic regression models in MATLAB 2024b using *fitglm* with a binomial distribution and logit link: one for awareness of the hearing loss–cognitive decline/dementia link and one for awareness of the hearing-aid claim (“yes” = 1; “no” = 0). “Not sure” responses (approximately 16% and 12%, respectively) were excluded from these binary contrasts but retained in descriptive comparisons (Tables S5 and S6). They may reflect uncertain recall or partial awareness and should not be treated as missing data or equated with “no”. Explanatory variables were education (coded 1–6), income (1–8), SCD, dementia concern (−2 to 2; DEconcern), SSQ, age, gender (0 = male; 1 = female), and population size. All were standardized before fitting. Variance inflation factors were below 1.4, indicating little collinearity among included variables (Belsley, Kuh, & Welsch, 1980; Chatterjee & Simonoff, 2012; O’brien, 2007). The model specification was:

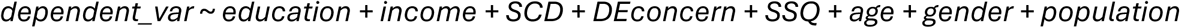

For Aim 2, two additional logistic regression models were estimated for participants who had heard about the link. The outcomes contrasted increased treatment inclination with no change, and greater openness in discussing hearing loss with no change (coded 1 and 0, respectively). Less treatment inclination (approximately 2.3%) and greater self-consciousness or worry (approximately 7%) were described but excluded from these binary models because they were infrequent. Models included the eight variables above (i.e., education, income, SCD, dementia concern, SSQ, age, gender, and population size) plus confidence in understanding the link and trust in healthcare professionals (each coded −2 to 2), all standardized. The variance inflation factor for all explanatory variables was below 1.6. These models characterize reported responses within the aware subgroup and cannot compare behavior between aware and unaware participants. The full model took the following form:

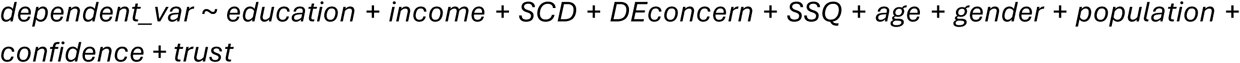

Models were exploratory and estimated mutually adjusted associations, not causal effects of individual variables (Westreich and Greenland, 2013). As an exploratory specification check, we varied covariate inclusion for variables significant in the full models using a multiverse analysis (Steegen et al., 2016; Hall et al., 2022; Mazei et al., 2025). These multiverse analyses describe sensitivity to adjustment choices (Supplementary Methods; Figures S3-S6). An association was considered robust if it showed a consistent direction and statistical significance across a large proportion of model specifications (Steegen et al., 2016). Correlations among explanatory variables are provided in Figure S1, and multiverse results in Figures S3–S6.

## Results

### Aim 1: Awareness of the hearing loss–dementia link and potential hearing-aid benefit

For the Canadian Viewpoint sample, about 27.5% of people had heard about the link between hearing loss and dementia/cognitive decline, whereas about 80% of health-engaged people had heard about the link (Figure 1A, Table 2). The percentage varied somewhat across provinces, but there were no statistically significant differences (p > 0.05; Figure 1A).

**Figure 1:**
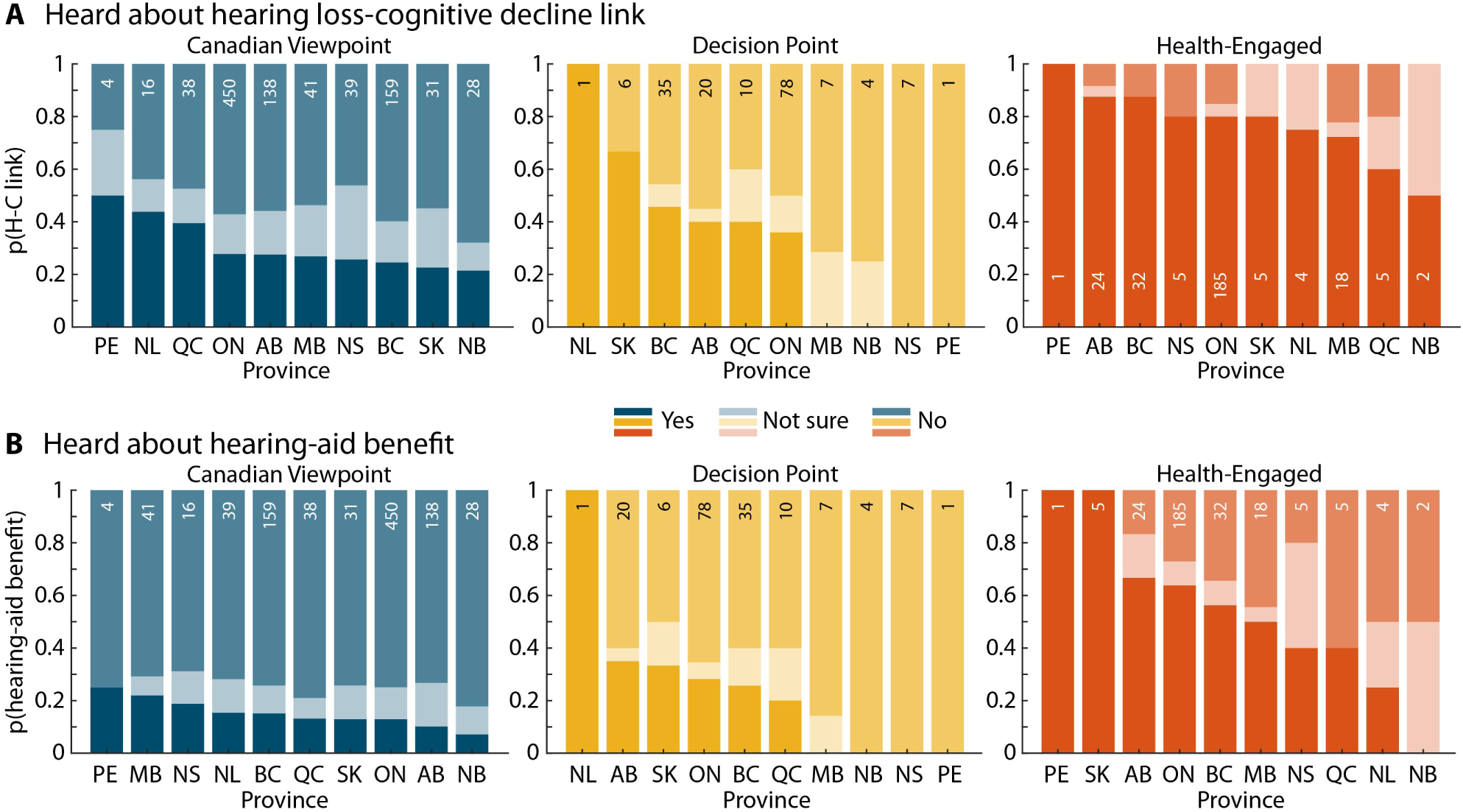
Proportion of individuals having heard about the hearing loss–cognitive decline/dementia link or hearing-aid benefit for different Canadian provinces. A: The proportion of having heard about the link between hearing loss and cognitive decline/dementia for each Canadian province. **B:** The proportion of having heard that hearing aids might help to prevent or reduce dementia/cognitive decline for each Canadian province. The small numbers within each bar indicate the number of participants from a specific province who answered the respective question. Data are split by the three recruitment sources: commercial panels (Canadian Viewpoint and Decision Point) and from health-engaged individuals. Provinces are sorted from highest to lowest proportion of ‘yes’ responses.

Higher education, greater dementia concern, older age, and female gender were associated with awareness of the link (Table 3; Figure 2A); their direction and statistical significance were consistent across the examined covariate specifications (Figure S3). Greater subjective cognitive decline was associated with lower awareness, mainly in models adjusting for dementia concern (Figures S2 and S3; a separate model testing the SCD × dementia concern interaction was not significant, p = 0.162). A model including all three recruitment samples yielded similar results (Table S1).

**Figure 2:**
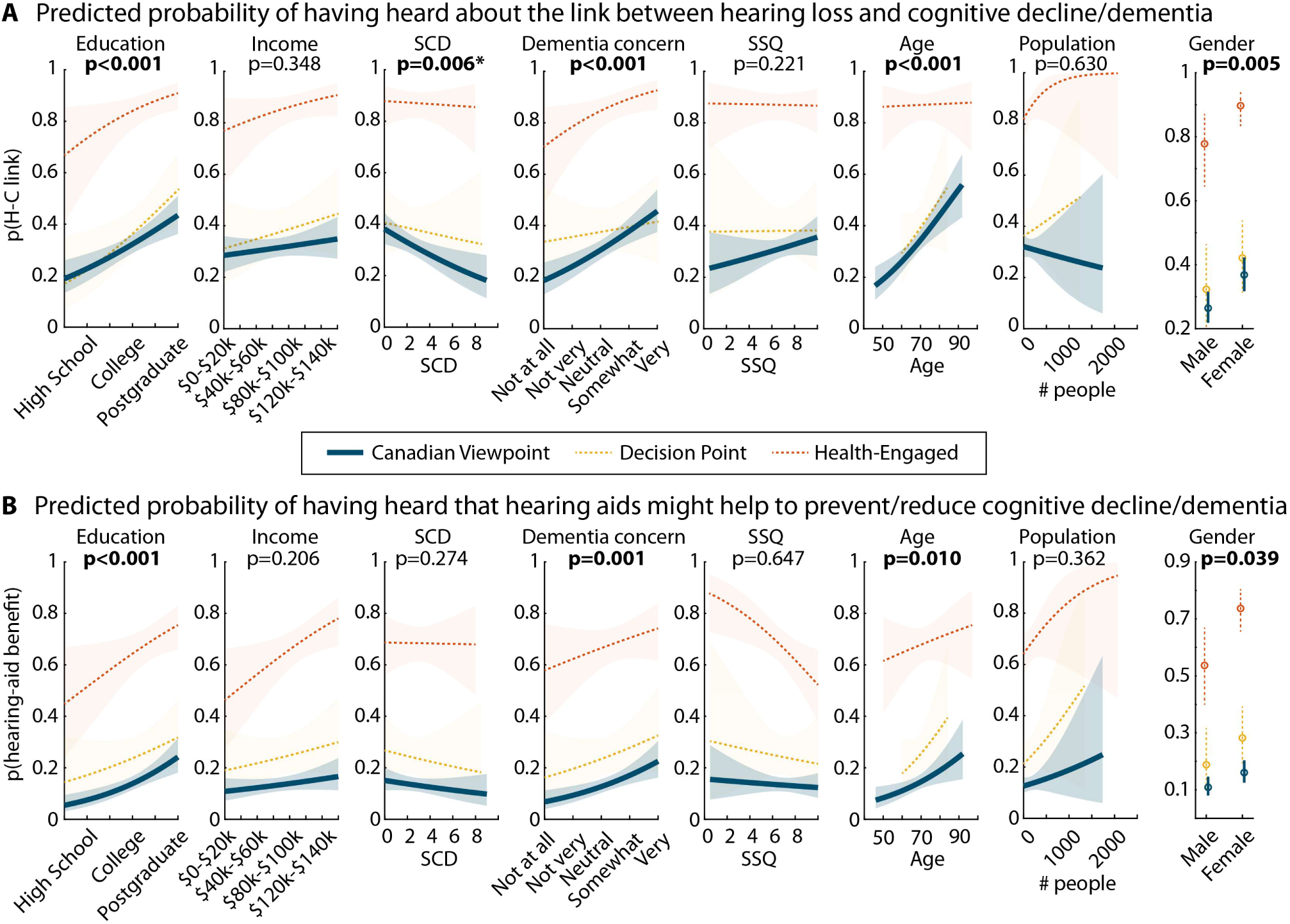
Model-predicted probabilities for each explanatory variable. A: Predicted probability of having heard about the link between hearing loss and cognitive decline/dementia. **B:** Predicted probability of having heard that hearing aids might help to prevent or reduce cognitive decline/dementia. Significance levels on the top of the graphs are provided for the Canadian Viewpoint sample. * indicates that the multiverse analysis suggests that this association depends on another explanatory variable. Specifically, the SCD effect was mostly conditional on dementia concern being accounted for in the model (Figure S2 and S3).

**Table 3:** Logistic regression model of the probability of a person having heard about the link between hearing loss and cognitive decline/dementia. * indicates that the multiverse analysis shows that this association depends on the inclusion of another explanatory variable: dementia concern (Figures S2 and S3). 710 individuals of the Canadian Viewpoint sample were included in the analysis after limiting the dependent variable to ‘yes’ and ‘no’ responses and listwise exclusion for missing data. Estimate – Estimated coefficient; SE – standard error of the estimate; Lower 95% and Upper 95% refer to the confidence interval of the odds ratio.

| Variable | Estimate | SE | Odds ratio | Lower 95% | Upper 95% | $t_{701}$ | p |
| --- | --- | --- | --- | --- | --- | --- | --- |
| Education | 0.341 | 0.0916 | 1.407 | 1.176 | 1.684 | 3.724 | **<0.001** |
| Income | 0.087 | 0.0922 | 1.091 | 0.910 | 1.307 | 0.939 | 0.348 |
| SCD | -0.274 | 0.0996 | 0.760 | 0.626 | 0.924 | -2.752 | **0.006\*** |
| DE concern | 0.375 | 0.0970 | 1.455 | 1.203 | 1.760 | 3.867 | **<0.001** |
| SSQ | 0.108 | 0.0882 | 1.114 | 0.937 | 1.324 | 1.222 | 0.222 |
| Age | 0.352 | 0.0875 | 1.422 | 1.198 | 1.688 | 4.023 | **<0.001** |
| Gender | 0.243 | 0.0865 | 1.275 | 1.076 | 1.510 | 2.806 | **0.005** |
| Population | -0.042 | 0.0866 | 0.959 | 0.810 | 1.137 | -0.481 | 0.630 |

In the Canadian Viewpoint sample, about 13.4% of people had heard that hearing aids might help to prevent or reduce cognitive decline/dementia, whereas 74.5% had not heard about it (Figure 1B). In contrast, 61.5% of health-engaged people had heard about the hearing-aid benefit, whereas 28% had not (Table 2). Differences across provinces were not significant (p > 0.05), except for the comparison between Manitoba and Alberta (p = 0.047 uncorrected; Figure 1B).

Higher education, greater dementia concern, older age, and female gender were also associated with awareness of the hearing-aid claim (Table 4; Figure 2B), although the age and gender associations were significant for many, but not all, covariate combinations (Figure S4). In the model including all three recruitment sources (Table S2), higher income and poorer subjective hearing were additionally associated with awareness.

**Table 4:** Logistic regression model of the probability of a person having heard that hearing aids might help to prevent or reduce cognitive decline/dementia. 744 individuals of the Canadian Viewpoint sample were included in the analysis after limiting the dependent variable to ‘yes’ and ‘no’ responses and listwise exclusion for missing data. Estimate – Estimated coefficient; SE – standard error of the estimate; Lower 95% and Upper 95% refer to the confidence interval of the odds ratio.

| Variable | Estimate | SE | Odds ratio | Lower 95% | Upper 95% | t<sub>735</sub> | p |
| --- | --- | --- | --- | --- | --- | --- | --- |
| Education | 0.484 | 0.1215 | 1.622 | 1.278 | 2.058 | 3.980 | **<0.001** |
| Income | 0.145 | 0.1144 | 1.156 | 0.923 | 1.446 | 1.264 | 0.206 |
| SCD | -0.134 | 0.1223 | 0.875 | 0.688 | 1.112 | -1.093 | 0.274 |
| DE concern | 0.398 | 0.1242 | 1.489 | 1.167 | 1.900 | 3.204 | **0.001** |
| SSQ | -0.0499 | 0.1090 | 0.951 | 0.768 | 1.178 | -0.457 | 0.647 |
| Age | 0.279 | 0.1088 | 1.321 | 1.068 | 1.635 | 2.561 | **0.010** |
| Gender | 0.225 | 0.1094 | 1.252 | 1.011 | 1.552 | 2.059 | **0.039** |
| Population | 0.085 | 0.0931 | 1.089 | 0.907 | 1.307 | 0.912 | 0.361 |

### Aim 2: Self-reported behavioral responses among participants aware of the link

The following analyses include only participants who had heard about the link between hearing loss and cognitive decline/dementia. For all three recruitment samples, the most common source of hearing about the link was news media, whereas other sources were reported less often (Figure 3A).

**Figure 3:**
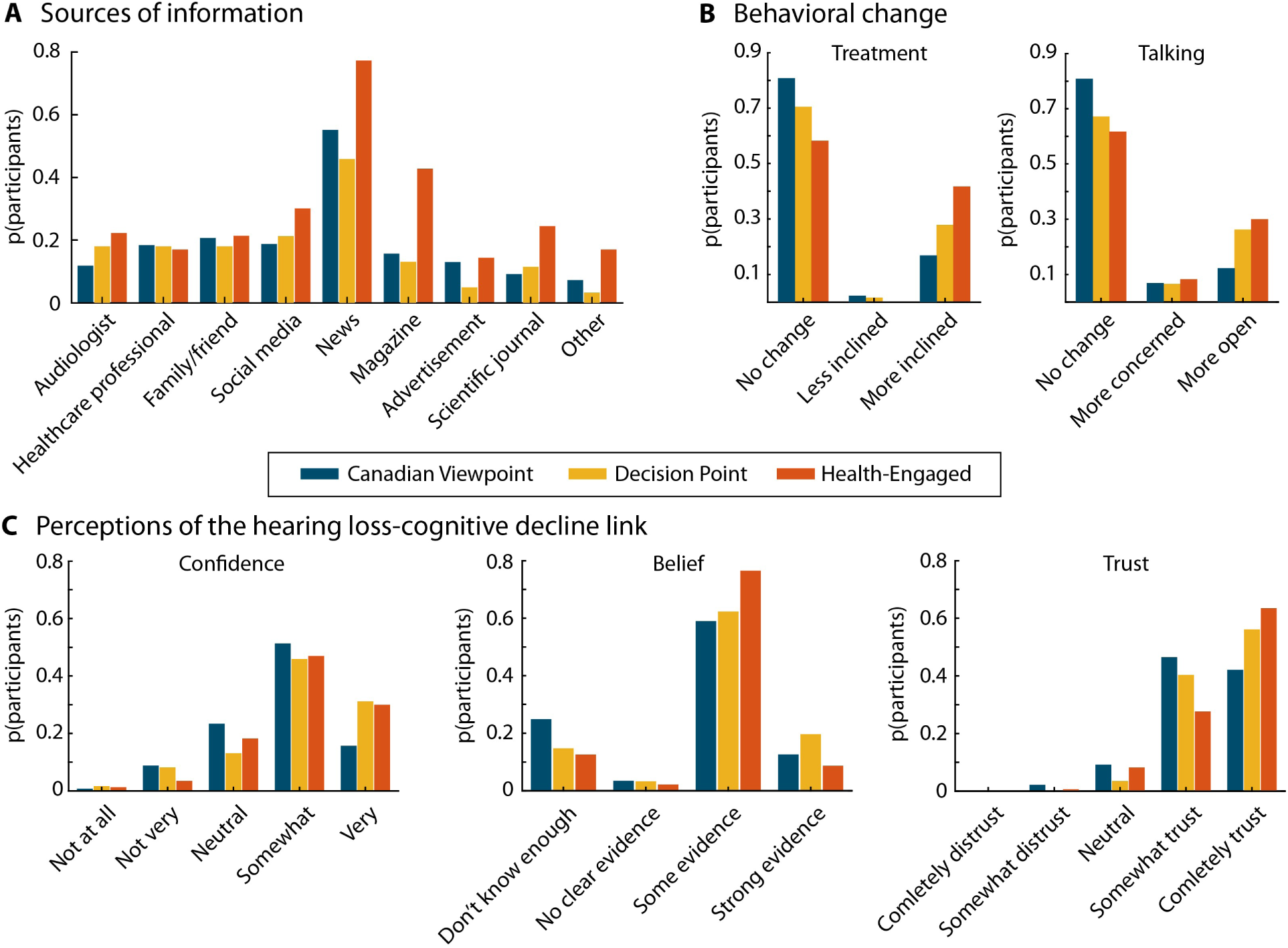
Overview of information sources, self-reported behavioral responses, and perceptions of the link between hearing loss and cognitive decline/dementia. A: Proportion of participants encountering different sources reporting on the link between hearing loss and cognitive decline/dementia (selection of more than one option was possible). **B:** Proportion of participants for different response categories for questions whether hearing about the link between hearing loss and cognitive decline/dementia changed the way they approach treatment (left) and talk about their hearing loss with others (right). **C:** Perceptions of the link between hearing loss and cognitive decline/dementia. Left: Confidence in their understanding of the link. Middle: Beliefs about the evidence of the link. Right: Trust of healthcare professionals when they talk about the link. Data are split by the three recruitment sources: commercial panels (Canadian Viewpoint and Decision Point) and from health-engaged individuals.

In the Canadian Viewpoint sample, 16.9% of aware participants reported being more inclined to seek or continue treatment and 12.3% reported talking more openly about their hearing loss as a result of having heard about the link. The numbers were 41.7% and 30%, respectively, for the health-engaged individuals (Table 2). A small proportion of participants reported being less inclined to seek/continue treatment (∼2%) or feeling more self-conscious or worried when talking about their hearing loss (∼7%), and these estimates were consistent across the three recruitment samples (Figure 3B).

Participants were mostly confident about their understanding of the link, mostly believed that there is some evidence that hearing loss may increase dementia risk, and mostly trust healthcare professionals when they talk about the link (Figure 3C).

Among aware Canadian Viewpoint participants, greater dementia concern, greater subjective cognitive decline, and higher confidence in understanding the link were associated with increased treatment inclination (Table 5; Figure 4A; multiverse analyses: Figure S5). Education, although associated with awareness, was not associated with this response. In the model including all three recruitment samples (Table S3), dementia concern and confidence remained associated with treatment inclination, whereas SCD was not significant.

**Figure 4:**
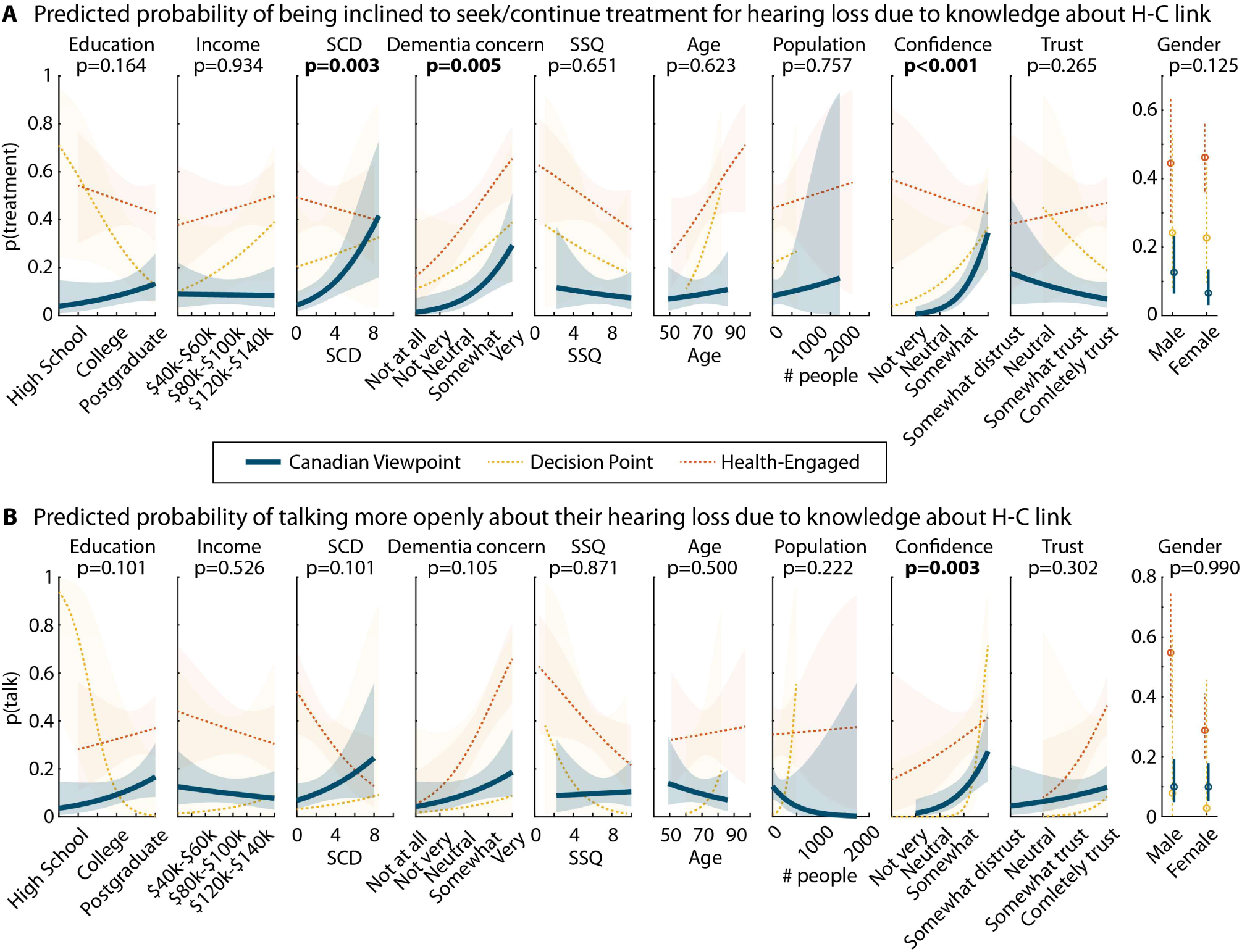
Model-predicted probabilities of self-reported behavioral responses for each explanatory variable. A: Predicted probability of a person being inclined to seek/continue treatment for hearing loss as a result of knowing about the link between hearing loss and cognitive decline/dementia (H-C link). **B:** Predicted probability of talking more openly about their hearing loss as a result of knowing about the link between hearing loss and cognitive decline/dementia. Significance levels on the top of the graphs are provided for the Canadian Viewpoint data.

**Table 5:** Logistic regression model of the probability of a person to be more inclined to seek/continue treatment for hearing loss. 197 individuals from the Canadian Viewpoint sample were included in the analysis after limiting the dependent variable to ‘More inclined’ and ‘No change’ responses and listwise exclusion for missing data. Also note that this analysis included only participants who indicated that they had heard about the link between hearing loss and cognitive decline/dementia. Estimate – Estimated coefficient; SE – standard error of the estimate; Lower 95% and Upper 95% refer to the confidence interval of the odds ratio.

| Variable | Estimate | SE | Odds ratio | Lower 95% | Upper 95% | t<sub>186</sub> | p |
| --- | --- | --- | --- | --- | --- | --- | --- |
| Education | 0.360 | 0.2584 | 1.433 | 0.864 | 2.377 | 1.392 | 0.164 |
| Income | -0.021 | 0.2505 | 0.979 | 0.599 | 1.601 | -0.083 | 0.934 |
| SCD | 0.722 | 0.2464 | 2.058 | 1.270 | 3.335 | 2.929 | **0.003** |
| DE concern | 0.954 | 0.3396 | 2.597 | 1.335 | 5.053 | 2.811 | **0.005** |
| SSQ | -0.107 | 0.2353 | 0.899 | 0.567 | 1.426 | -0.453 | 0.651 |
| Age | 0.114 | 0.2327 | 1.121 | 0.711 | 1.769 | 0.491 | 0.623 |
| Gender | -0.356 | 0.2319 | 0.701 | 0.445 | 1.104 | -1.534 | 0.125 |
| Population | 0.074 | 0.2406 | 1.077 | 0.672 | 1.726 | 0.309 | 0.757 |
| Confidence | 1.175 | 0.3001 | 3.239 | 1.799 | 5.831 | 3.916 | **<0.001** |
| Trust | -0.259 | 0.2325 | 0.772 | 0.489 | 1.217 | -1.115 | 0.265 |

Higher confidence in understanding the link was the only variable associated with talking more openly about one’s hearing loss as a result of having heard about the link (Table 6; Figure 4B; multiverse analyses: Figure S6). In pooled-source analyses, confidence remained associated with openness, alongside greater dementia concern, poorer subjective hearing, and greater trust in healthcare professionals (Table S4).

**Table 6:** Logistic regression model of the probability of a person to talk more openly about their hearing loss as a result of having heard about the link between hearing loss and cognitive decline/dementia. 189 individuals from the Canadian Viewpoint sample were included in the analysis after limiting the dependent variable to ‘More open’ and ‘No change’ responses and listwise exclusion for missing data. Also note that this analysis included only participants who indicated that they had heard about the link between hearing loss and cognitive decline/dementia. Estimate – Estimated coefficient; SE – standard error of the estimate; Lower 95% and Upper 95% refer to the confidence interval of the odds ratio.

| Variable | Estimate | SE | Odds ratio | Lower 95% | Upper 95% | $t_{178}$ | p |
| --- | --- | --- | --- | --- | --- | --- | --- |
| Education | 0.457 | 0.2784 | 1.579 | 0.915 | 2.725 | 1.641 | 0.101 |
| Income | -0.157 | 0.2467 | 0.855 | 0.527 | 1.387 | -0.635 | 0.526 |
| SCD | 0.394 | 0.2400 | 1.482 | 0.926 | 2.373 | 1.640 | 0.101 |
| DE concern | 0.460 | 0.2836 | 1.583 | 0.908 | 2.761 | 1.620 | 0.105 |
| SSQ | 0.038 | 0.2362 | 1.039 | 0.654 | 1.651 | 0.163 | 0.871 |
| Age | -0.173 | 0.2290 | 0.841 | 0.537 | 1.318 | -0.756 | 0.450 |
| Gender | -0.003 | 0.2299 | 0.997 | 0.635 | 1.565 | -0.013 | 0.990 |
| Population | -0.426 | 0.3494 | 0.653 | 0.329 | 1.295 | -1.220 | 0.222 |
| Confidence | 0.862 | 0.2928 | 2.367 | 1.333 | 4.202 | 2.942 | **0.003** |
| Trust | 0.254 | 0.2460 | 1.289 | 0.796 | 2.088 | 1.032 | 0.302 |

## Discussion

This survey in middle-to older-aged Canadians examined awareness of, and self-reported behavioral responses to, the message that hearing loss is linked to cognitive decline/dementia. About 28% of people had heard of the link and 13% had heard of a possible cognitive benefit of hearing aids. Awareness was higher among individuals with more education, greater dementia concerns, older age, and females. Among aware participants, 17% reported increased treatment inclination and 12% greater openness about hearing loss. These responses were associated with dementia concern, subjective cognitive decline, and confidence in understanding the link rather than with education. Both awareness and these responses were more common in the health-engaged convenience sample. These data provide evidence of how a societal narrative that hearing health is cognitive health reaches and potentially affects behavior of community-dwelling middle-to older-aged Canadians.

### Awareness of the hearing loss–dementia link and potential hearing-aid benefit

Fewer than 30% of panel participants had heard of the link between hearing loss and cognitive decline/dementia, and fewer still (13%) had heard of a potential cognitive benefit of hearing aids. This pattern suggests that the broad idea that hearing loss and cognitive decline/dementia are linked has entered public discourse more readily than the more specific, and currently more uncertain, proposition that treating hearing loss might mitigate dementia risk.

Our estimate falls within the range reported when hearing loss has been included in dementia-literacy surveys: only 4% of Icelandic adults (Jónsdóttir et al., 2022), 18% of Norwegian adults (Kjelvik et al., 2022), 28% of Australian adults (Nagel et al., 2021), and 35% of Irish adults (Dukelow et al., 2023) endorsed hearing loss as dementia risk factor, whereas recognition was higher (59%) in a recent Bulgarian sample (Lazarova and Petrova-Antonova, 2025). Qualitative research shows that the public tends to associate dementia prevention mainly with lifestyle domains such as exercise, diet, and mental or social activity, with sensory health receiving little spontaneous attention (Curran et al., 2021). Nevertheless, awareness in our health-engaged sample (80%) shows that the message can permeate substantially further among people connected to health and aging networks.

Awareness of the hearing-loss–dementia link followed socioeconomic and demographic gradients similar to those reported for dementia literacy more generally, where knowledge tends to be higher among people with more education and, in some studies, among females and older adults (Smith et al., 2014; Cahill et al., 2015; Heger et al., 2019; Milani et al., 2019; Pacifico et al., 2022; Korkmaz Yaylagül et al., 2024; Lazarova and Petrova-Antonova, 2025), although these patterns are not consistent across countries (Cahill et al., 2015). The COM-B lens suggests that education likely increases both the opportunity to encounter health information and the capability to process it. People with lower socioeconomic status and health literacy, who are also more likely to experience untreated hearing loss and other dementia risk factors, are thus less likely to have encountered the message, echoing calls for dementia-prevention campaigns to pay particular attention to these groups (Van Asbroeck et al., 2021; Paauw et al., 2024; Eddy-Lacey et al., 2025). Population size of the area of residence, our proxy for urbanicity, was not associated with awareness, but we could not assess neighborhood characteristics more directly.

Greater concern about dementia was also associated with awareness of both the hearing loss–dementia link and the potential hearing-aid benefit, consistent with evidence that people who are more worried about dementia are especially attentive to information about risk and prevention (Ashworth et al., 2021; Curran et al., 2021). Our cross-sectional design cannot determine whether dementia concern leads people to seek out and retain such information, or whether exposure to the message increases concern. Nonetheless, the association suggests that communication about hearing and cognitive decline/dementia is intertwined with broader emotional responses to cognitive aging, which may shape how information is received and interpreted.

Participants most frequently reported learning about the link from news sources, mirroring the media interest in hearing and dementia in Canada (Timson, 2017; Taylor, 2018; McKnight, 2019; La Grassa, 2023; Jiang, 2024a). News coverage can raise awareness quickly, but it may also oversimplify the evidence or imply that hearing loss causes dementia (Blustein et al., 2020; Pichora-Fuller, 2023; Dawes and Munro, 2024; Pichora-Fuller and Mick, 2024). Nevertheless, most aware participants endorsed the view that there is some, rather than strong, evidence linking hearing loss to cognitive decline (Figure 3C), a cautious understanding that aligns with ongoing debates about the link and with the modest effects of hearing interventions reported to date (Lin et al., 2023; Dawes and Munro, 2024; Pichora-Fuller and Mick, 2024).

### Self-reported behavioral responses to the hearing loss–dementia link

Among respondents who had heard about the hearing loss–dementia link, about 17% reported being more inclined to seek or continue treatment, and about 12% reported talking more openly about their hearing loss. These numbers were more than twice as high in our health-engaged subsample. For some people, framing hearing care as relevant to brain health may thus prompt a re-evaluation of hearing problems, which is relevant given longstanding challenges of delayed help-seeking, low hearing-aid uptake, and concealment of hearing difficulties (Chien and Lin, 2013; Davidson et al., 2021; Reed et al., 2023). It is important to note, however, that these figures reflect retrospective self-reports, not observed behavior, and whether they translate into hearing assessments or sustained hearing-aid use is unknown. Moreover, because only aware participants were asked these questions, our data cannot show whether awareness itself is associated with more help-seeking than among people who have not heard the message.

Yet, the majority of aware participants did not report behavior change. This echoes broader risk-communication research showing that awareness of a risk factor is not sufficient to alter behavior (Marteau et al., 2012; Kelly and Barker, 2016). Hearing-aid uptake and use are shaped by the degree of hearing loss, perceived hearing handicap, cost, stigma, social support, and prior hearing-aid experience (Knudsen et al., 2010), which likely predate and overlap with the variables measured here. Moreover, some participants may already have been using hearing aids, may not have hearing difficulties that call for treatment, may perceive barriers that outweigh perceived benefits, or may view the evidence as not strong enough to justify action, which would be a realistic reading of the mixed evidence for cognitive benefits of hearing interventions (Sanders et al., 2021; Yang et al., 2022; Lin et al., 2023). Because the survey did not assess hearing-aid use or audiometric hearing status, we could not distinguish these possibilities.

Reported treatment inclination was more likely among people with lower subjective cognitive function and greater dementia concern, consistent with work in other domains showing that health worries can be a strong driver of behavioral change (Chapman and Coups, 2006; Ferrer et al., 2012; Ferrer and Klein, 2015; Kapoor and Tagat, 2021). In contrast, although people with higher education were more likely to know about the link, they were not more likely to report a behavior change (Figures 2 and 4). This is in line with the COM-B framework (Michie et al., 2011), highlighting greater opportunities to encounter a message differ from factors that provide the motivation to act on it. Campaigns that focus solely on the number of people knowing about the link may overestimate their behavioral impact (Kelly and Barker, 2016).

Confidence in understanding the link was associated with both self-reported treatment-related and communication-related change. This is consistent with health-behavior models highlighting self-efficacy and coherent cognitive representations of health risks (Bandura, 1977; Rosenstock et al., 1988; Leventhal et al., 2016; Hagger and Orbell, 2022), and suggests that fostering clear understanding, not only exposure, may matter for whether a message is acted upon. However, individuals who act on the message may also subsequently feel more confident about their knowledge of the link. Audiologists, primary-care physicians, and other clinicians whom most participants trusted, are well placed to translate complex, evolving evidence into balanced, individualized discussions.

A small subset of participants reported potentially negative responses, such that they felt less inclined to seek treatment (2%) or more self-conscious and worried when talking about their hearing loss (7%). Although numerically small, these responses highlight concerns that emphasizing dementia risk may exacerbate anxiety or intensify stigma rather than empower action (Pichora-Fuller, 2023; Dawes and Munro, 2024; Pichora-Fuller and Mick, 2024). Our findings thus do not imply that the goal should be to maximize awareness of the hearing loss– dementia link, but to facilitate accurate understanding through communication that conveys what the evidence does and does not show.

### Limitations

The cross-sectional design and retrospective self-report of behavioral responses preclude causal inferences. We cannot determine whether the message changed participants’ intentions or whether people who are already more proactive about their hearing and brain health are also more likely to encounter and endorse such messages. Longitudinal or experimental designs, perhaps embedded within hearing or cognitive-health programs, could clarify temporal relationships and link message exposure to observed behavior.

Participants were recruited through commercial online panels and health-related organizations, had to be proficient in English, and needed internet access. People without internet access, with lower literacy, or who are not proficient in English, including many immigrants and French-speaking Canadians, are therefore likely underrepresented, and our estimates should not be generalized to the Canadian middle-to older-aged population as a whole. Panel members may also be more health-conscious or technologically engaged than non-members. The large differences between recruitment sources illustrate how strongly selection can affect prevalence estimates. Probability-based sampling or integrating questions into large national health surveys in both official languages, would help assess generalizability.

Awareness and responses were measured using single items with categorical response options, and hearing and cognition were assessed via a brief SSQ (Gatehouse and Noble, 2004; Noble et al., 2013) and a subjective cognitive decline scale (Gifford et al., 2015) rather than audiometric and neuropsychological measures. The survey also did not assess hearing-aid ownership, recent hearing assessments, or other known determinants of hearing-aid use, which limits our ability to account for factors that could modify the associations observed.

Lastly, survey items referred to both “dementia” and “cognitive decline,” which differ in clinical meaning and emotional salience. Reported awareness and responses could therefore reflect engagement with a broadly framed hearing–cognition narrative rather than a precise understanding of dementia risk.

## Conclusions

In a large online sample of middle-to older-aged Canadians, fewer than one in three had heard that hearing loss may be linked to cognitive decline/dementia, and awareness followed socioeconomic and demographic gradients. Among those who had heard about the link, most reported no change in how they approach hearing care, and reported responses were associated with dementia concern and confidence in understanding the link rather than with education. Awareness of the link alone thus appears to be a weak lever for behavior change. Given the uncertainty about the causal link, communication efforts may be better aimed at accurate, balanced understanding across diverse groups than at maximizing awareness.

## Data Availability

All data produced are available online at https://osf.io/f4d9t/ (upon publication).

## Acknowledgments

We thank the Canadian Hard of Hearing Association for their input and support. Language editing (grammar correction, sentence shortening, and paragraph condensing) was assisted by Claude Opus 5 (Anthropic; https://claude.ai). The authors reviewed all revisions and take full responsibility for the final text.

## Author contributions

**SB:** Conceptualization, methodology, formal analysis, investigation, data curation, writing - original draft, writing - review and editing; **SLM:** Conceptualization, methodology, investigation, writing - review and editing; **MB:** formal analysis, writing - review and editing; **BH:** Conceptualization, methodology, formal analysis, investigation, writing - original draft, writing - review and editing, visualization, supervision, project administration, funding acquisition.

## Statements and Declarations

The authors have no conflicts or competing interests.

## Data availability

The survey data used in this study are available in the repository of the Open Science Framework under https://osf.io/f4d9t/ (upon publication).

