## Supplementary Materials for "Public awareness of the hearing loss-dementia link and self-reported behavioral changes in middle-to older-aged Canadians"

### Supplementary methods

#### English-language proficiency screen and attention checks

The survey began with a brief English-language proficiency test to ensure participants could understand and complete the questionnaire. The test consisted of two items ("Fill in the blank: His eyes were ...... bad that he couldn't read the number plate of the car in front." response options [such, so, too, very]; and "Select the correct response to the following phrase: Can I park here?" response options ["Sorry, I did that.", "It's the same place.", "Only for half an hour."]), and participants who did not answer both questions correctly did not proceed to the main survey.

To identify inattentive respondents, the survey also included several attention-check items (e.g., "Please select 'Never' to confirm that you have read this question carefully"), which were used during data cleaning to exclude invalid respondents

#### Data Cleaning

Across all recruitment sources, a total of 480 participants were excluded prior to analysis, leading to a final sample of 1404 datasets across recruitment sources. Fifty-two participants were removed for not meeting the eligibility criteria of being a native or proficient English speaker and a resident of Canada (Canadian Viewpoint: 33; Decision Point Research: 16; Health-Engaged: 3), 19 for not providing consent (CV: 18; DP: 0; HE: 1), and 96 for failing the English proficiency test (CV: 70; DP: 21; HE: 5). An additional 218 participants were excluded for incomplete or abandoned surveys (CV: 130; DP: 21; HE: 67), 5 for surveys completed in under five minutes (CV: 3; DP: 1; HE: 1), and 90 for failing one or more of the embedded attention-check item (CV: 56; DP: 15; HE: 19).

After these exclusions, the final sample sizes were 948 for Canadian Viewpoint, 170 for Decision Point, and 286 for the Health-Engaged sample. Because participants were able to provide free-text responses for age, gender, and province of residence, these variables were systematically re-coded to provide consistency and usability for analysis. Gender responses were standardized (e.g., responses such as "a woman" and typos such as "mail" were recoded to "female" and "male," respectively). Age entries provided as words (e.g., "seventy-eight") were converted to numeric values, and province names were abbreviated using standard two-letter postal abbreviations (e.g., "Ontario" became "ON"). Minor corrections were also made to ensure consistent spelling and capitalization across all entries. These steps were applied only to correct obvious inconsistencies and did not alter the meaning of participants' responses.

Several participants selected 'other' to describe their education idiosyncratically. Forty-two of these responses were deemed close to the pre-determined categories used in the survey and were thus recoded (CV: 25; DP: 6; HE: 11). Examples include the recoding of participants' responses 'college for trade school', 'technical college', 'community college', 'business college', etc. to the survey category ‘College Degree (2 years)’; ‘military college’ to ‘Bachelor's Degree’; and ‘some post secondary in college’, ‘some post secondary’, ‘some college’ to the survey category ‘Some University’. The recoding enabled us to use the respondents’ data for analysis rather than excluding them entirely due to list-wise exclusion in the regression models (note that education effects reported below did not depend on the inclusion of participants for whom education was recoded).

### Supplementary results

#### Statistical modeling with the inclusion of recruitment samples

We fitted the same statistical models as reported in the main article with the addition of the categorical covariate ‘recruitment source’ (Canadian Viewpoint, Decision Point, Health-Engaged). The results reported in the main article are mirrored and, in some cases, extended when all data samples are included in the model (Tables S1–S4). Overall, these additional analyses highlight the robustness of the results reported in the main article.

| Variable | Estimate | SE | Odds ratio | Lower 95% | Upper 95% | $T_{1061}$ | p |
|---|---|---|---|---|---|---|---|
| Education | 0.382 | 0.0779 | 1.466 | 1.258 | 1.708 | 4.909 | **<0.001** |
| Income | 0.129 | 0.0784 | 1.137 | 0.975 | 1.326 | 1.641 | 0.101 |
| SCD | -0.235 | 0.0831 | 0.790 | 0.672 | 0.930 | -2.835 | **0.005*** |
| DE concern | 0.353 | 0.0811 | 1.423 | 1.214 | 1.668 | 4.353 | **<0.001** |
| SSQ | 0.063 | 0.0748 | 1.066 | 0.920 | 1.234 | 0.848 | 0.397 |
| Age | 0.294 | 0.0751 | 1.342 | 1.158 | 1.555 | 3.914 | **<0.001** |
| Gender | 0.269 | 0.0731 | 1.309 | 1.134 | 1.511 | 3.680 | **<0.001** |
| Population | 0.038 | 0.0716 | 1.039 | 0.903 | 1.195 | 0.531 | 0.595 |
| Source DP | 0.037 | 0.2021 | 1.037 | 0.698 | 1.542 | 0.181 | 0.856 |
| Source HE | 1.846 | 0.2133 | 6.337 | 4.172 | 9.626 | 8.659 | **<0.001** |

**Table S1:** Logistic regression model of the probability of a person having heard the link between hearing loss and cognitive decline/dementia. * indicates that the multiverse analysis shows that this association depends on the inclusion of another explanatory variable: dementia concern (Figures S2 and S3. 1072 individuals were included in the analysis after limiting the dependent variable to ‘yes’ and ‘no’ responses and listwise exclusion for missing data. Estimate – Estimated coefficient; SE – standard error of the estimate; Lower 95% and Upper 95% refer to the confidence interval of the odds ratio. Effects of Source DP (Decision Point) and Source HE (Health-Engaged) reflect the contrast relative to Canadian Viewpoint data.

| Variable | Estimate | SE | Odds ratio | Lower 95% | Upper 95% | $T_{1091}$ | p |
|---|---|---|---|---|---|---|---|

| | | | | | | | |
|---|---|---|---|---|---|---|---|
| Education | 0.423 | 0.0902 | 1.526 | 1.279 | 1.821 | 4.685 | **<0.001** |
| Income | 0.212 | 0.0881 | 1.236 | 1.040 | 1.469 | 2.409 | **0.016** |
| SCD | -0.124 | 0.0925 | 0.883 | 0.737 | 1.059 | -1.345 | 0.179 |
| DE concern | 0.328 | 0.0925 | 1.389 | 1.158 | 1.664 | 3.549 | **<0.001** |
| SSQ | -0.179 | 0.0807 | 0.836 | 0.714 | 0.979 | -2.217 | **0.027** |
| Age | 0.227 | 0.0843 | 1.254 | 1.063 | 1.480 | 2.689 | **0.007** |
| Gender | 0.284 | 0.0842 | 1.329 | 1.127 | 1.567 | 3.375 | **<0.001** |
| Population | 0.133 | 0.0735 | 1.142 | 0.988 | 1.319 | 1.803 | 0.071 |
| Source DP | 0.427 | 0.2267 | 1.532 | 0.983 | 2.389 | 1.883 | 0.060 |
| Source HE | 1.831 | 0.1960 | 6.238 | 4.248 | 9.160 | 9.338 | **<0.001** |

**Table S2:** Logistic regression model of the probability of a person having heard that hearing aids might help to prevent or reduce cognitive decline/dementia. 1102 individuals were included in the analysis after limiting the dependent variable to 'yes' and 'no' responses and listwise exclusion for missing data. Estimate – Estimated coefficient; SE – standard error of the estimate; Lower 95% and Upper 95% refer to the confidence interval of the odds ratio. Effects of Source DP (Decision Point) and Source HE (Health-Engaged) reflect the contrast relative to Canadian Viewpoint data.

| Variable | Estimate | SE | Odds ratio | Lower 95% | Upper 95% | $T_{374}$ | p |
|---|---|---|---|---|---|---|---|
| Education | -0.016 | 0.1383 | 0.984 | 0.750 | 1.290 | -0.118 | 0.906 |
| Income | 0.126 | 0.1445 | 1.134 | 0.854 | 1.506 | 0.872 | 0.383 |
| SCD | 0.204 | 0.1410 | 1.226 | 0.930 | 1.617 | 1.447 | 0.148 |
| DE concern | 0.626 | 0.1605 | 1.869 | 1.365 | 2.560 | 3.898 | **<0.001** |
| SSQ | -0.239 | 0.1291 | 0.788 | 0.612 | 1.015 | -1.848 | 0.065 |
| Age | 0.235 | 0.1343 | 1.265 | 0.972 | 1.647 | 1.752 | 0.080 |
| Gender | -0.133 | 0.1341 | 0.875 | 0.673 | 1.139 | -0.993 | 0.321 |
| Population | 0.074 | 0.1203 | 1.077 | 0.850 | 1.363 | 0.613 | 0.540 |
| Confidence | 0.400 | 0.1480 | 1.492 | 1.117 | 1.994 | 2.705 | **0.007** |
| Trust | -0.085 | 0.1438 | 0.918 | 0.693 | 1.217 | -0.593 | 0.553 |
| Source DP | 0.621 | 0.4016 | 1.860 | 0.847 | 4.087 | 1.546 | 0.122 |
| Source HE | 1.094 | 0.3102 | 2.985 | 1.625 | 5.482 | 3.525 | **<0.001** |

**Table S3:** Logistic regression model of the probability of a person to be more inclined to seek/continue treatment for hearing loss. 387 individuals were included in the analysis after limiting the dependent variable to 'yes, more inclined' and 'no change' responses and listwise exclusion for missing data. Estimate – Estimated coefficient; SE – standard error of the estimate; Lower 95% and Upper 95% refer to the confidence interval of the odds ratio. Effects of Source DP (Decision Point) and Source HE (Health-Engaged) reflect the contrast relative to Canadian Viewpoint data.

| Variable | Estimate | SE | Odds ratio | Lower 95% | Upper 95% | $T_{351}$ | p |
|---|---|---|---|---|---|---|---|
| Education | 0.005 | 0.1489 | 1.005 | 0.750 | 1.345 | 0.032 | 0.975 |
| Income | -0.074 | 0.1529 | 0.929 | 0.688 | 1.254 | -0.482 | 0.630 |
| SCD | -0.099 | 0.1504 | 0.906 | 0.674 | 1.216 | -0.658 | 0.510 |

| | | | | | | | |
|---|---|---|---|---|---|---|---|
| DE concern | 0.580 | 0.1621 | 1.785 | 1.299 | 2.453 | 3.575 | **<0.001** |
| SSQ | -0.277 | 0.1349 | 0.758 | 0.582 | 0.988 | -2.051 | **0.040** |
| Age | 0.019 | 0.1429 | 1.020 | 0.771 | 1.349 | 0.136 | 0.892 |
| Gender | -0.128 | 0.1462 | 0.880 | 0.661 | 1.172 | -0.875 | 0.382 |
| Population | 0.048 | 0.1378 | 1.050 | 0.801 | 1.375 | 0.352 | 0.725 |
| Confidence | 0.599 | 0.1735 | 1.820 | 1.296 | 2.558 | 3.452 | **0.001** |
| Trust | 0.430 | 0.1750 | 1.537 | 1.091 | 2.166 | 2.457 | **0.014** |
| Source DP | 0.820 | 0.4144 | 2.271 | 1.008 | 5.115 | 1.979 | **0.048** |
| Source HE | 0.946 | 0.3396 | 2.576 | 1.324 | 5.013 | 2.787 | **0.005** |

**Table S4:** Logistic regression model of the probability of a person to talk more openly about their hearing loss as a result of having heard about the link between hearing loss and cognitive decline/dementia. 364 individuals were included in the analysis after limiting the dependent variable to 'yes, more open' and 'no change' responses and listwise exclusion for missing data. Estimate – Estimated coefficient; SE – standard error of the estimate; Lower 95% and Upper 95% refer to the confidence interval of the odds ratio. Effects of Source DP (Decision Point) and Source HE (Health-Engaged) reflect the contrast relative to Canadian Viewpoint data.

### Characteristics of 'not sure' respondents for Aim 1 outcome variables

The two outcome variables of Aim 1 included 'not sure' as a response category, in addition to 'no' and 'yes'. Participants who responded 'not sure' were not included in the main analyses because it is ambiguous whether this response reflects partial awareness, uncertain recall, or a distinct response type. Tables S5 and S6 compare these respondents with 'no' and 'yes' respondents. For the question about the hearing loss–dementia link (Table S5), 'not sure' respondents resembled 'yes' respondents in education (both higher than 'no' respondents), but reported more subjective cognitive decline and poorer subjective hearing than 'yes' respondents. For the question about a potential hearing-aid benefit (Table S6), 'not sure' respondents were more educated than 'no' respondents and reported less dementia concern than 'yes' respondents. 'Not sure' respondents thus do not map clearly onto either binary category.

| **Variable** | **'no'** | **'not sure'** | **'yes'** | **no vs not sure** | **not sure vs yes** | **no vs yes** |
|---|---|---|---|---|---|---|
| N | 532 | 155 | 261 | - | - | - |
| Population | 1244 | 1332 | 1200 | 0.590 | 0.469 | 0.741 |
| Age | 66.17 | 66.74 | 68.29 | 0.484 | 0.083 | **0.001** |
| Gender | 0.456 | 0.526 | 0.586 | 0.124 | 0.232 | **<0.001** |
| Education | 3.641 | 4.1275 | 4.079 | **<0.001** | 0.734 | **<0.001** |
| Income | 4.228 | 4.110 | 4.409 | 0.530 | 0.149 | 0.241 |
| SCD | 2.770 | 3.055 | 2.347 | 0.211 | **0.003** | **0.020** |
| SSQ | 6.909 | 6.616 | 7.159 | 0.073 | **0.002** | 0.057 |
| Dementia concern | 0.066 | 0.135 | 0.241 | 0.509 | 0.357 | **0.044** |

**Table S5: Descriptive data and comparisons between participants responding differently to the question whether they have heard about the link between hearing loss and cognitive decline/dementia.** The three columns of the left side reflect mean responses (gender reflects the proportion of females) for participants responding ‘no’, ‘not sure’, or ‘yes’ for the Canadian Viewpoint sample (N=948). The three columns on the right side reflect significance levels for contrasts between different responders using an independent samples t-test (continuous) or chi-square test (categorical). See Table 2 in the main article and related text for descriptions about the numerical coding of the other variables.

| **Variable** | **‘no’** | **‘not sure’** | **‘yes’** | **no vs not sure** | **not sure vs yes** | **no vs yes** |
|---|---|---|---|---|---|---|
| N | 706 | 115 | 127 | - | - | - |
| Population | 1219 | 1175 | 1465 | 0.795 | 0.244 | 0.160 |
| Age | 66.52 | 67.30 | 68.28 | 0.383 | 0.380 | **0.040** |
| Gender | 0.487 | 0.522 | 0.575 | 0.492 | 0.407 | 0.069 |
| Education | 3.718 | 4.046 | 4.339 | **0.027** | 0.098 | **<0.001** |
| Income | 4.181 | 4.407 | 4.551 | 0.275 | 0.584 | 0.060 |
| SCD | 2.751 | 2.413 | 2.673 | 0.170 | 0.384 | 0.739 |
| SSQ | 6.936 | 6.900 | 6.919 | 0.838 | 0.933 | 0.919 |
| Dementia concern | 0.103 | -0.026 | 0.386 | 0.266 | **0.005** | **0.010** |

**Table S6: Descriptive data and comparisons between participants responding differently to the question whether they have heard that hearing aids may prevent/reduce dementia/cognitive decline.** The three columns of the left side reflect mean responses (gender reflects the proportion of females) for participants responding ‘no’, ‘not sure’, or ‘yes’ for the Canadian Viewpoint sample (N=948). The three columns on the right side reflect significance levels for contrasts between different responders using an independent samples t-test (continuous) or chi-square test (categorical). See Table 2 in the main article and related text for descriptions about the numerical coding of the other variables.

### Correlation matrix for explanatory variables

The correlation between individual explanatory variables was calculated to assess the relationship among variables used in the logistic regression models described in the main article. Correlations are shown in Figure S1.

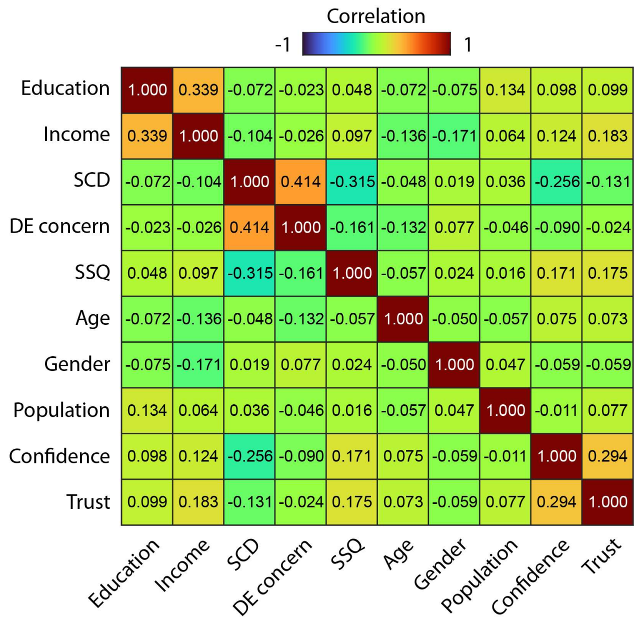


**Figure S1: Correlation matrix for explanatory variables.**

### SCD effect conditional on dementia concern

The logistic regression modeling the probability of having heard about the link between hearing loss and cognitive decline/dementia showed a negative association with SCD that was significant mainly when the variable 'dementia concern' was also included in the model (Figure S3). Figure S2 depicts the predicted probability of having heard about the link as a function of SCD, separately for each of the five levels of dementia concern.

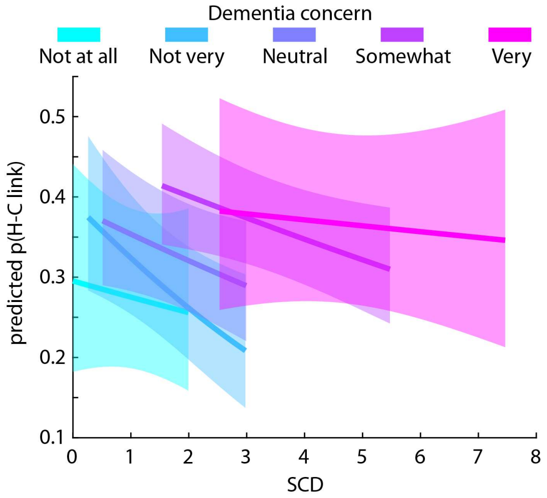


**Figure S2: Probability of having heard about the link between hearing loss and dementia/cognitive decline, depending on SCD and dementia concern.** The SCD × dementia concern interaction was not significant (p = 0.162). The SCD range for each dementia concern level was limited to the 25th and 75th percentile to highlight the correlation between SCD and dementia concern.

### Multiverse analyses

For explanatory variables significant in the full models, we fitted logistic regressions retaining the variable of interest while varying all combinations of the other covariates (Hall et al., 2022; Mazei et al., 2025; Steegen et al., 2016; references in the main article). Figures S3–S6 display coefficients and statistical significance across these specifications. This exploratory analysis describes dependence on adjustment choices, including the SCD association's dependence on adjustment for dementia concern. Specifications are not equally justified causal models, and consistency across them does not establish freedom from confounding, multiplicity, or selection bias. Only variables significant in the full models were examined, so these displays do not provide an independent validation of the initial findings.

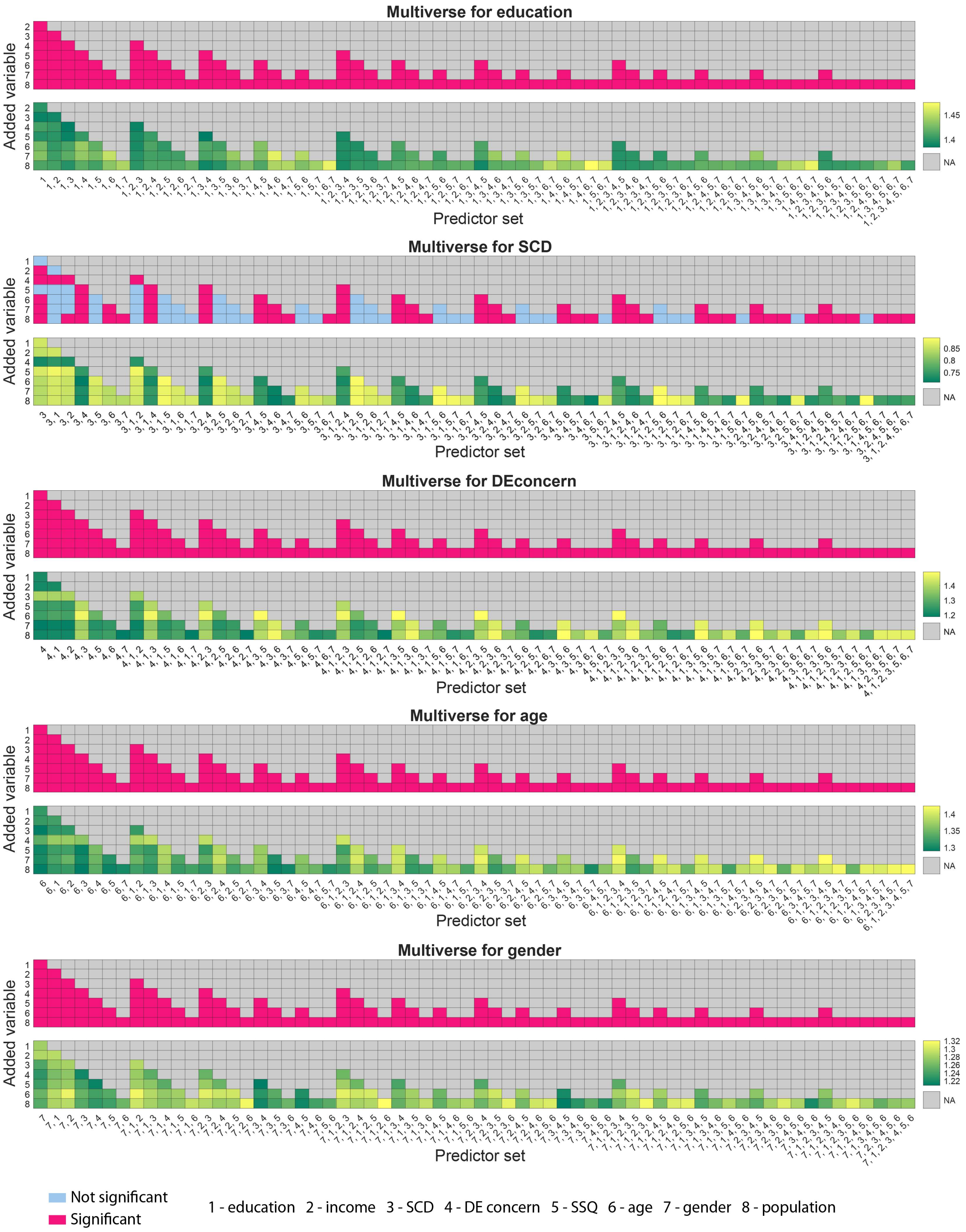
Multiverse for education
Added variable
Predictor set
Multiverse for SCD
Added variable
Predictor set
Multiverse for DEconcern
Added variable
Predictor set
Multiverse for age
Added variable
Predictor set
Multiverse for gender
Added variable
Predictor set
NA
Not significant
Significant
1 - education 2 - income 3 - SCD 4 - DE concern 5 - SSQ 6 - age 7 - gender 8 - population

**Figure S3: Multiverse analyses for the model of the probability of a person having heard the link between hearing loss and cognitive decline/dementia.** For each explanatory variable of interest, logistic regression models were calculated that included the specific variable of interest and varied the inclusion of the remaining variables across all possible combinations. The top graph in each pair of graphs shows whether the variable of interest was significant (red) or not (blue) given the inclusion of other variables in the model. The bottom graph of each pair shows the estimated coefficient of the variable of interest given the inclusion of other variables in the model. The rows and columns jointly reflect the variables included in the model. For example, for the education multiverse (top pair of graphs), column 1 and row 3 reflects the significance level and estimated coefficient for education from the model including variables ‘education’ and ‘DE concern’ (dementia concern), whereas column 2 and row 6 reflects the significance level and estimated coefficient for education from the model including variables ‘education’, ‘income’, and ‘gender’. Numbers on the x- and y- axis indicate specific variables.

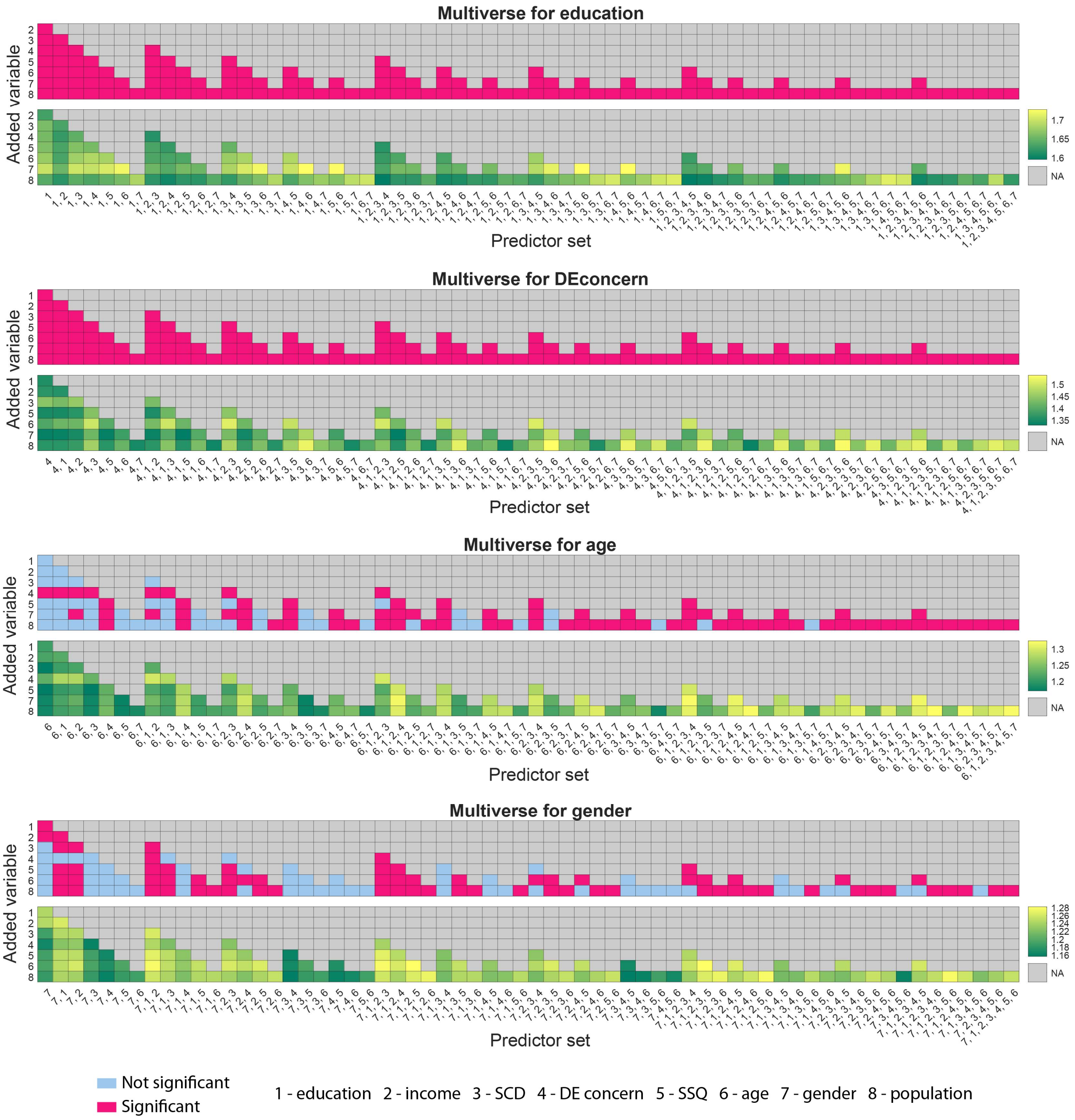


**Figure S4: Multiverse analyses for the model of the probability of a person having heard that hearing aids might help to prevent or reduce cognitive decline/dementia.** For each explanatory variable of interest, logistic regression models were calculated that included the specific variable of interest and varied the inclusion of the remaining variables across all possible combinations. The top graph in each pair of graphs shows whether the variable of interest was significant (red) or not (blue) given the inclusion of other variables in the model. The bottom graph of each pair shows the estimated coefficient of the variable of interest given the inclusion of other variables in the model. The rows and columns jointly reflect the variables included in the model. For example, for the education multiverse (top pair of graphs), column 1 and row 3 reflects the significance level and estimated coefficient for education from the model including variables ‘education’ and ‘DE concern’ (dementia concern), whereas column 2 and row 6 reflects the significance level and estimated coefficient for education from the model including variables ‘education’, ‘income’, and ‘gender’. Numbers on the x- and y- axis indicate specific variables.

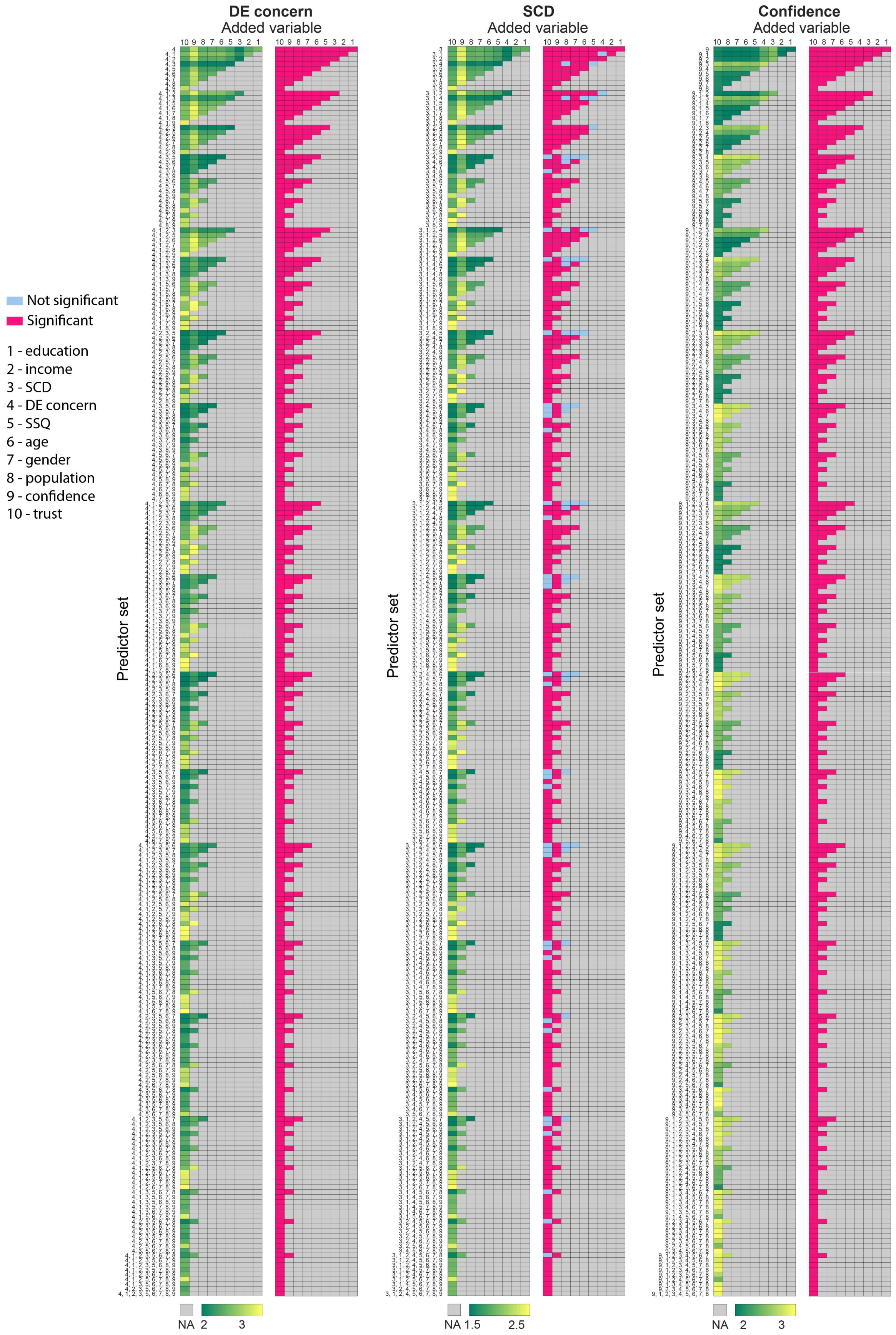
DE concern
Added variable
SCD
Added variable
Confidence
Added variable
Not significant
Significant
1 - education
2 - income
3 - SCD
4 - DE concern
5 - SSQ
6 - age
7 - gender
8 - population
9 - confidence
10 - trust
Predictor set
NA 2 3
NA 1.5 2.5
NA 2 3

**Figure S5: Multiverse analyses for the model of the probability of a person to be more inclined to seek/continue treatment for hearing loss.** For each explanatory variable of interest, logistic regression models were calculated that included the specific variable of interest and varied the inclusion of the remaining variables across all possible combinations. The right graph in each pair of graphs shows whether the variable of interest was significant (red) or not (blue) given the inclusion of other variables in the model. The left graph of each pair shows the estimated coefficient of the variable of interest given the inclusion of other variables in the model. The rows and columns jointly reflect the variables included in the model. For example, for the SCD multiverse (middle pair of graphs), row 1 and column 4 reflects the significance level and estimated coefficient for SCD from the model including variables 'SCD' and 'gender'. Numbers on the x- and y- axis indicate specific variables.

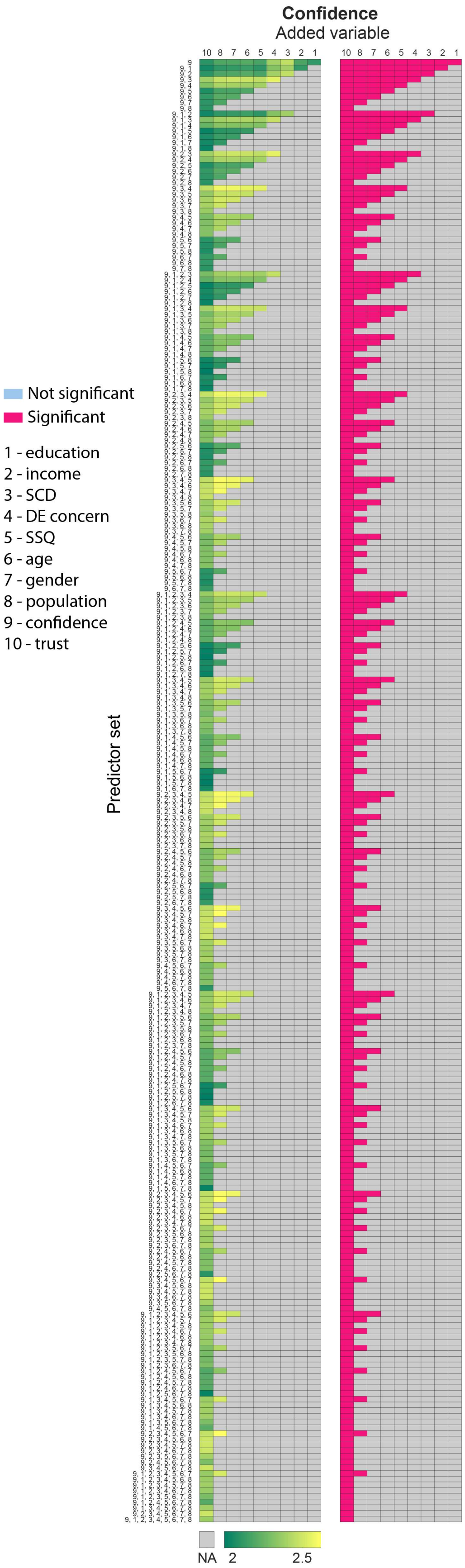


**Figure S6: Multiverse analyses for the model of the probability of a person to talk more openly about their hearing loss.** For the explanatory variable of interest, logistic regression models were calculated that included the specific variable of interest and varied the inclusion of the remaining variables across all possible combinations. The right graph of the pair of graphs shows whether the variable of interest was significant (red) or not (blue) given the inclusion of other variables in the model. The left graph of the pair shows the estimated coefficient of the variable of interest given the inclusion of other variables in the model. The rows and columns jointly reflect the variables included in the model. For example, row 1 and column 4 reflects the significance level and estimated coefficient for Confidence from the model including variables ‘Confidence and ‘gender’. Numbers on the x- and y- axis indicate specific variables.
